# Developing a blood cell-based diagnostic test for myalgic encephalomyelitis/chronic fatigue syndrome using peripheral blood mononuclear cells

**DOI:** 10.1101/2023.03.18.23286575

**Authors:** Jiabao Xu, Tiffany Lodge, Caroline Kingdon, James W L Strong, John Maclennan, Eliana Lacerda, Slawomir Kujawski, Pawel Zalewski, Wei E. Huang, Karl J. Morten

## Abstract

Myalgic encephalomyelitis/chronic fatigue syndrome (ME/CFS) is characterized by debilitating fatigue that profoundly impacts patients’ lives. Diagnosis of ME/CFS remains challenging, with most patients relying on self-report, questionnaires, and subjective measures to receive a diagnosis, and many never receiving a clear diagnosis at all. In this study, we utilized a single-cell Raman platform and artificial intelligence to analyze blood cells from 98 human subjects, including 61 ME/CFS patients of varying disease severity and 37 healthy and disease controls. Our results demonstrate that Raman profiles of blood cells can distinguish between healthy individuals, disease controls, and ME/CFS patients with high accuracy (91%), and can further differentiate between mild, moderate, and severe ME/CFS patients (84%). Additionally, we identified specific Raman peaks that correlate with ME/CFS phenotypes and have the potential to provide insights into biological changes and support the development of new therapeutics. This study presents a promising approach for aiding in the diagnosis and management of ME/CFS, and could be extended to other unexplained chronic diseases such as long COVID and post-treatment Lyme disease syndrome, which share many of the same symptoms as ME/CFS.

## Introduction

Myalgic encephalomyelitis/chronic fatigue syndrome (ME/CFS) is a complex disease with unknown aetiology. With the hallmark of disease being fatigue, ME/CFS has a wide range of symptoms that vary between patients and can fluctuate over time. These symptoms often include post-exertional malaise (PEM), cognitive dysfunction, sleep disturbances, orthostatic intolerance, abnormal thermal regulation, myalgia, photosensitivity, and neuro-immuno-endocrine dysfunction (1), all of which result in a significant reduction in patients’ quality of life. Although the precise trigger(s) for ME/CFS remain(s) undetermined, it has been proposed that infectious agents play a role in initiating the disorder. Recently, many COVID-19 patients have developed Long Covid or Long Hauler syndrome (2–4), which may be classified as ME/CFS if unresolved after six months and if it meets the IOM criteria for ME/CFS, which mandate a PEM component (5). There are over 250,000 ME/CFS cases reported in the UK, with this number likely to increase due to Long Covid (6–8). Around 70% of ME/CFS patients are female. Females have a stronger response to infection or vaccination than males, and differences in sex hormones are proposed to play a role in the prevalence of autoimmune diseases, including ME/CFS (9). Additionally, the microbiome may be essential, as different microbiota have been associated with different hormonal profiles in males and females (10). Evidence also suggests abnormal brain function in ME/CFS, with microglial activation being studied directly or indirectly linked to metabolic changes and inflammation (11).

ME/CFS lacks a single sensitive and specific diagnostic test (12), making the development of a simple test with the potential for early diagnosis a critical goal. Early diagnosis would enable patients to manage their conditions more effectively, potentially leading to new discoveries in disease pathways and treatment development. Given that most ME/CFS cases are identified via symptoms and questionnaires, the exclusion of alternative diagnoses is vital. Blood-based biomarkers may prove useful in quickly and accurately diagnosing ME/CFS by supplementing current sets of indicators measured during routine medical check-ups. Furthermore, blood-derived markers may help differentiate similar disorders, such as ME/CFS, multiple sclerosis (MS), fibromyalgia, chronic Lyme disease, and Long Covid (7). Blood draws could also provide longitudinal insights into the treatment response for ME/CFS and the onset of more severe symptoms. Overall, developing a blood-based, single, objective test would be a significant step toward enhancing the diagnosis and management of ME/CFS.

Peripheral blood mononuclear cells (PBMCs) and muscle biopsies obtained from ME/CFS patients exhibited altered mitochondrial function, indicating a difference in energetic function when compared to non-fatigued controls (13–15). Research by Missailidis et al. discovered that when lymphocytes from ME/CFS patients were immortalized, they generated cell lines with very different energetic properties (16). As ME/CFS may have a systemic energy issue, studying PBMCs may provide a good model for understanding the pathology affecting other organ systems. With the evidence suggesting differences in the blood cell fractions from ME/CFS patients, we hypothesized that single-cell analysis of PBMCs might reveal differences in ME/CFS compared to healthy and other disease groups. In this study, we have included MS patients as a disease control group. ME/CFS and MS are two distinct conditions with many similar clinical symptoms. One of the key differences between the conditions is that MS is associated with clear pathological changes in the brain and is considered by the medical profession as a real illness (17) while ME/CFS is still viewed with scepticism by many with no effective treatment options or clear pathology.

Raman spectroscopy is a non-invasive and label-free approach to probe molecular vibrations in a sample, and when combined with confocal microscopy, it can interrogate individual cells (18). A single-cell Raman spectrum (SCRS) is a phenotypic fingerprint of all biomolecules in that cell and could potentially differentiate between various cell types and give insights into underlying biology (18). Our previous pilot study demonstrated that a comparison of SCRS could distinguish between ME/CFS patients and healthy controls, and identified a potential PBMC biomarker for ME/CFS (19). Here, we built on our pilot study and further assessed the diagnostic potential of a blood-based platform using single-cell Raman spectroscopy and state-of-the-art ensemble learning classification models to discriminate ME/CFS from two control groups. We also evaluated the capability of the approach to differentiating between different ME/CFS disease severity groups, including mild, moderate and severe. With PBMC being an easily accessible target, we believe that Raman spectroscopy combined with advanced artificial intelligence could offer an affordable and non-invasive screening tool for ME/CFS when the condition is first identified.

## Results

### Study design of human subjects and clinical characteristics

Our previous pilot study on 10 individuals illustrated the capability of single-cell Raman spectroscopy and machine learning on approaches in finding specific biomarkers in PBMCs of ME/CFS patients (19). We now expanded our approach to a larger cohort as a blood-based Raman spectroscopic diagnostic test at the single-cell level. Table S1 summarises the key characteristics of the ME/CFS cohort and the healthy (HC) and disease (MS) controls involved in this study. In total, 98 human subjects were involved, including 61 ME/CFS and 37 controls (HC, n = 16; MS, n = 21), based on their clinical profiles. The ME/CFS cohort was further divided into Severe (n = 21), Moderate (n = 15) and Mild (n = 25).

All patients in the fatigue groups (ME/CFS and MS) had fatigue for at least six months that was not relieved by rest. All MS patients had a MS diagnosis given by an NHS consultant (20) and those recruited were a mixture of relapsing, remitting and progressive forms. Those with ME/CFS required a previous medical diagnosis of ME/CFS. To be accepted as a participant with ME/CFS, potential donors must meet either the Canadian Consensus Criteria or CDC-1994 criteria; many fulfil both. The assessment process for compliance with study criteria includes baseline questionnaires about symptoms, a clinical assessment performed by a clinical member of the research team, and urinalysis screening and baseline blood tests, which are used to exclude alternative diagnoses. Detailed questioning of potential participants with ME/CFS enables their disease to be classified according to different case definitions. ME/CFS cases were categorized as mild if they had an SF-36 Physical Function (PF) score greater than 25; moderate if the PF score was below 25; and severe if they were house- or bed-bound, with all patients identified as suffering with PEM.

Fatigue levels of the ME/CFS, MS patients and healthy controls were measured by clinical measures of Fatigue Severity Scale (FSS) (21). The General Health Questionnaire (GHQ) (22) was used primarily as a screening tool to exclude patients suffering from severe depression and has already been used in this capacity in a previous ME/CFS study using Fukuda 1994 criteria (23). The median FSS values could distinguish ME/CFS and MS patients from healthy controls; ME/CFS: 59 (range 44–63); MS: 54 (range 16–63); HC 17 (range 11–37) (Table S1). However, the FSS evaluation could not easily separate the ME/CFS from the MS patients nor between different ME/CFS severities (p = 0.07). Using additional clinical measures recorded by the UK ME/CFS biobank during recruitment, we investigated their utility in discriminating ME/CFS from MS disease controls and from healthy controls, producing a symptom burden assessment (Table S2). Symptom feature inclusion was determined by calculating the relative mean ordinal intensity for each variable to provide sufficient detail to rank order (as opposed to a median derived integer with categorical representation) symptoms, allowing for selective inclusion with a >1.5-fold difference between groups (severe ME compared to MS). Figure 1 summarizes the symptoms into a heatmap showing the top 28 clinical variables (Figure 1). A clear difference was observed when comparing the HC with the ME/CFS and MS patients. An increase in severity of symptoms was seen when moving from mild to severe ME/CFS patients. The most significant measures as assessed by Fischer’s Exact Test were calculated based on the severe ME vs MS comparison, with the Benjamini-Hochberg (BH) procedure applied to adjust for multiple comparisons (*, p < 0.05).

**Figure 1.**
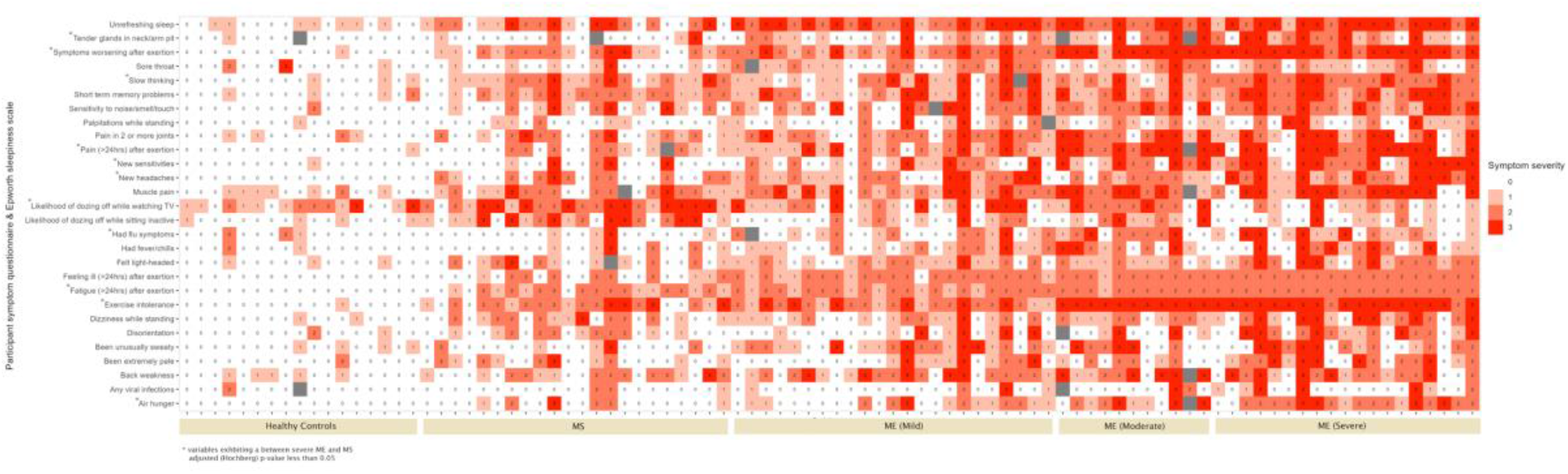
Cohort clinical characteristics. Symptom presence and intensity were determined for 63 variables obtained from the UKMEB symptoms assessment. Responses were recorded on an ordinal 4-point scale, with 0 indicating “absent”, 1 indicating “mild”, 2 indicating “moderate” and 3 indicating “severe”; grey boxes indicate missing data. Category inclusion was determined by calculating the relative mean ordinal weight/intensity for each variable, with a between-group (severe ME compared to MS as the reference group) fold differentiation ≥1.5 mandated for analytical inclusion. Additionally, Fischer’s Exact Test was calculated for severe ME vs MS comparison, with the Benjamini-Hochberg (BH) procedure applied to adjust for multiple comparisons (sig. p < 0.05).

### SCRS differentiate the different cohorts

We first used a simple approach examining mitochondrial oxidative phosphorylation in viable frozen PBMCs from 41 out of the 98 subjects. A previous report by Tomas et al. has shown a difference in whole-cell mitochondrial respiration, consistent with a deficiency in cellular energetics associated with mitochondrial dysfunction or substrate flux feeding into the TCA cycle and mitochondrial respiratory chain (15). However, this assay was difficult to reproduce; Missailidis et al. failed to reproduce this finding in PBMCs but did find differences in immortalised lymphocytes (24). In our study, cell viability following thawing was between 70–85% with a noticeable drop in viability following 24 hr in culture. Mitochondrial respiration was measured in 5-mM glucose media with rates measured over 1–2 hr. No difference was observed in rates of mitochondrial respiration between ME/CFS patients, MS patients and healthy controls (Figure S1A). When ME/CFS patients were divided into severe, moderate and mild patients, no difference was observed (Figure S1B). This demonstrated that mitochondrial function assessment of PBMCs using an oxygen consumption assay on cryopreserved frozen samples failed to discriminate disease cohorts, and will be challenging to develop as a robust diagnostic approach.

As a simple mitochondrial function assessment failed to differentiate different cohorts, we then sought to use single-cell Raman spectroscopy to obtain whole single-cell molecular profiles on the same samples. PBMCs of 98 individuals were measured by single-cell Raman spectroscopy with single-cell Raman spectra (SCRS) were acquired on single PBMCs. In total, we obtained 14600 spectra from 2155 single cells. All Raman measurements were blinded in this study. Figure 2A presents the averaged SCRS of single PBMCs from the HC (number of cells = 410), ME/CFS (number of cells = 1151) and MS cohorts (number of cells = 594) at the fingerprint region (300–1800 cm^−1^). The fingerprint region contains most information from intrinsic molecular vibrations inside a cell and therefore, can be regarded as a “fingerprint” of one cell. The cell-to-cell fluctuation was represented as the standard deviation at each wavenumber, drawn as the shaded area in Figure 2A. The sum of fluctuation was 21.7%, 19.7% and 17.1% in HC, ME/CFS and MS, respectively. The cells in the ME/CFS cohorts differed more than those in MS cohorts in terms of their metabolic profiles, probably because of a higher number of subjects involved and the broad range of symptom severities from mild, moderate and severe. Similarly, averaged SCRS for ME/CFS subgroups of Mild, Moderate and Severe are illustrated in Figure 2B. Figure 2C demonstrates the difference of spectra between ME/CFS and HC, as well as between MS and HC. In addition to differing from healthy controls, the two disease cohorts also showed a number of spectral differences from each other.

**Figure 2.**
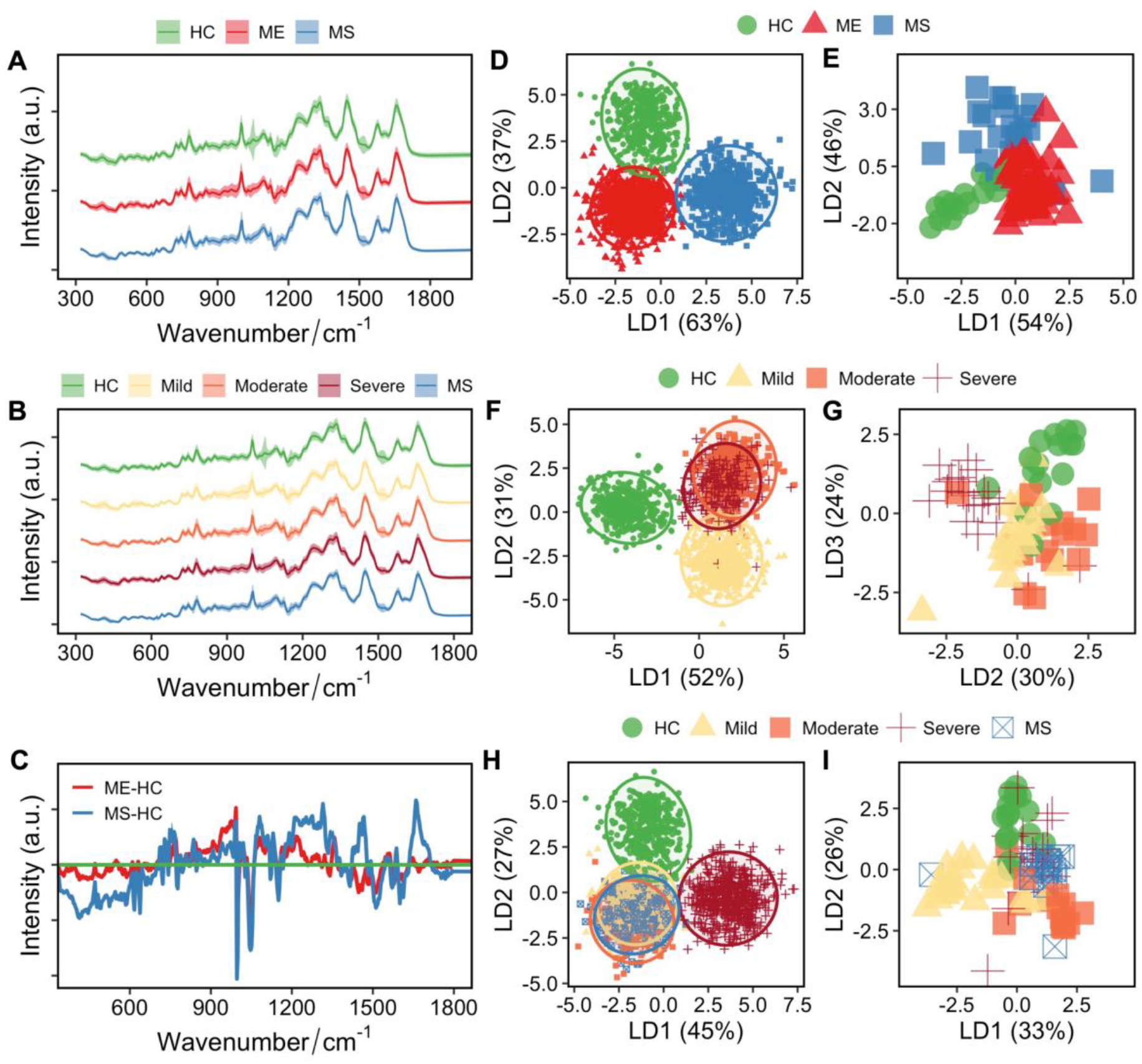
SCRS differ among different cohorts. Averaged Raman spectra of 2155 single cells obtained from 98 individual subjects, separating into **(A)** three groups of HCs, ME and MS, or **(B)** five groups of HCs, Mild ME, Moderate ME, Severe ME and MS. **(C)** Differences between spectra of ME and HC and MS and HC. Raman spectra from each group was shifted in intensity to aid visualisation and the intensity is expressed in arbitrary unit (a.u.). LDA clustering was used to visualise separations among three groups of HC, ME and MS at the single-cell level **(D)** and at the individual level **(E**), four groups of HC and different ME groups (Mild, Moderate, and Severe) at the single-cell level **(F)** and at the individual level **(G**), and five groups of HC, different ME groups (Mild, Moderate, and Severe) and MS at the single-cell level **(H)** and at the individual level **(I**).

Supervised linear discriminant analysis (LDA) was used to reduce the noise and the high dimensionality of Raman spectra due to the presence of 1019 Raman wavenumbers (spectral range from 319 to 3401 cm^−1^; spectral step size of ∼3 cm^−1^). An LDA plot of all SCRS of 2155 cells along LD1 and LD2 illustrates three clearly separable clusters representing HC, ME/CFS and MS (Figure 2D). The LD1, which explains 63% of the data variance, separates the MS cohort from the healthy controls and the ME/CFS cohort, while LD2 explains the remaining 37% variance and separates the ME/CFS cohort from the others. In addition to visualization using single cells, LDA visualization was then conducted at the individual level showing 98 subjects as three distinct clusters for HC, ME, and MS (Figure 2E). To compare HC with ME/CFS cohorts only, LDA was performed on four groups: HC and the 3 ME/CFS subgroups. Intriguingly, an LDA plot of all SCRS again separates the HC and ME subgroups where the axis along LD1 (52% of the variance) distinguishes all ME cohorts from the healthy controls and LD2 (31% of the variance) reveals the division of disease clusters of mild and groups of moderate and severe (Figure 2F). Averaging SCRS to individuals nicely elucidates differences between HC subjects and ME disease groups with different severities of mild, moderate, and severe (Figure 2G). An LDA with all five groups shows distinctive clusters of HC and severe ME, while interestingly, an overlap of mild and moderate ME/CFS and MS (Figure 2H). LDA of the five groups on the subject level also displays the separation of different subgroups (Figure 2I). Overall, data from the three-group, four-group, and five-group LDA shows the underlying differentiating power within the SCRS for fatigue diseases like ME/CFS and MS.

We then evaluated possible confounding factors that could have influenced the LDA separations of the different cohorts, including sex, body mass index (BMI), age, disease duration, types and total counts of medications and supplements, as well as the freezing duration and processing time for each sample (Table S3). Pearson correlation coefficients were obtained from pairs of variables; the highest correlation with regard to LD1 values was −0.24 (Medication Class Opiate Present) and the highest correlation against LD2 was −0.27 (Medication Class Tricyclic Or Mirtazapine Present), both of which are well below the definition for high correlation (>0.75). The low correlation between classification values and all potential confounding factors illustrates the robustness of the Raman dataset to overlook baseline differences and highlight disease effects.

### Quantification of biomolecules in different cohorts

The LDA was also used as a feature selection tool to find the most significant Raman peak features contributing to group separation. Quantification at the significant peaks with respect to different groups of HCs, ME/CFS and MS highlighted the different biological components in cells of healthy individuals and fatigue patients. Top Raman peak features selected based on LDA clustering were quantified at single wavenumbers for HC, ME/CFS and MS groups, respectively (Table S4).

Following on from our pilot study finding phenylalanine as a potential biomarker in PBMCs of ME/CFS patients, we highlighted the relative quantification of aromatic amino acids (AAAs) among cohorts, namely tryptophan (Figure 3A), tyrosine (Figure 3B), and phenylalanine (Figure 3C) by integrating and quantifying the associated Raman bands. A universal increase in tryptophan and tyrosine in cellular PBMCs was observed in all disease cohorts, including in all subgroups of ME/CFS and the MS group (Figure 3A and 3B). Significance was found in all groups compared to the healthy controls except for the mild ME/CFS subgroup. Quantification of intracellular phenylalanine, on the other hand, suggests metabolic subtypes existing in the ME patients, with the moderate and severe groups having significantly reduced phenylalanine and the mild ME and MS having increased levels relative to controls. Besides amino acids, altered lipid metabolism was also found (Figure 3D, 3E and 3F). All the disease cohorts had significantly elevated glycerol levels compared to the healthy controls. Inconsistent alterations in unsaturated lipids were observed with elevated levels in the mild ME and the MS cohorts and decreased levels in the moderate and severe ME patients (Figure 3E), suggesting a difference in the lipidic profiles of different cohorts. Reduced cholesterol and cholesteryl esters were observed, especially evident in the MS cohort. Biomolecules related to energy fuelling also demonstrated differences among cohorts. Glycogen levels were significantly reduced in the mild and severe ME cohort, as well as in the MS group. Glucose quantification showed a decrease in all ME subgroups and the MS cohort had the lowest glucose accumulation.

**Figure 3.**
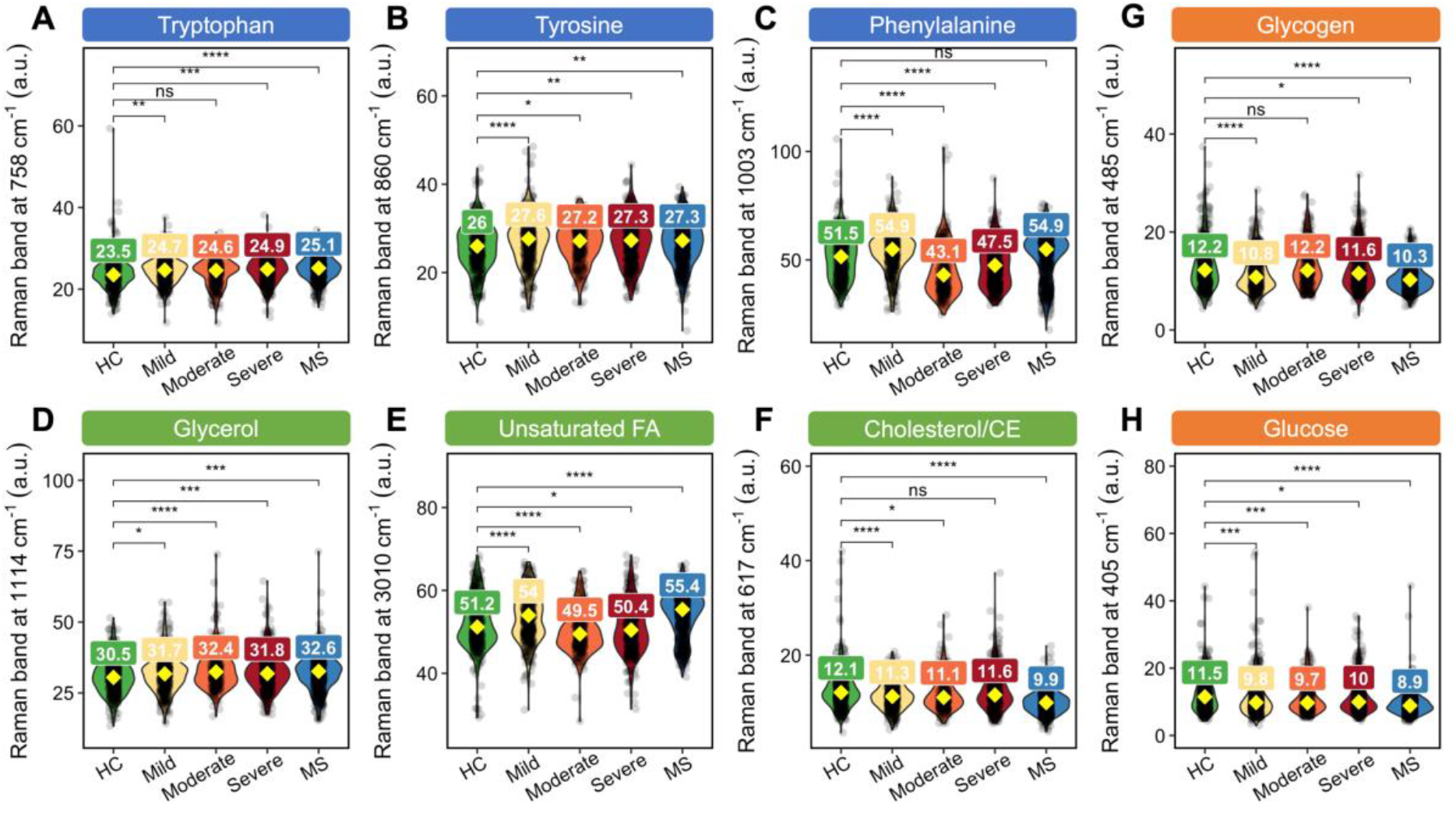
Relative quantification of biomolecules in PBMCs of HC, ME (mild, moderate and severe) and MS cohorts, related to aromatic amino acids (AAAs) of **(A)** tryptophan at 758 cm^−1^, **(B)** tyrosine at 860 cm^−1^ and **(C)** phenylalanine at 1003 cm^−1^, lipid metabolism of **(D)** glycerol at 1114 cm^−1^, **(E)** unsaturated fatty acids (FA) at 3010 cm^−1^ and **(F)** cholesterol/cholesteryl esters (CE) at 617 cm^−1^, and energy metabolism of **(G)** glycogen at 485 cm^−1^ and **(H)** glucose at 405 cm^−1^. The quantification results were represented as box plots and sample mean of each disease group was compared with healthy control (HC) by using Welch’s two sample t-test for unequal variance (ns: not significant; **: p < 0.01; ***: p < 0.001; ****: p < 0.0001).

### A cell-based diagnostic test using SCRS of PBMCs and an ensemble learner

As LDA has already demonstrated its capability to well separate different disease cohorts, we then sought to evaluate its performance as a classifier on diagnosis by training a classification model with 80% of the spectral data and testing the model performance by using the remaining 20% as an independent test set. The accuracy of classifying five classes of MS, Severe ME, Moderate ME, Mild ME and HC was 54.8% on the train set and 47.1% on the test set (Table S5). In order to further improve the differentiation power of the model, ensemble learning was employed. Ensemble learning is a machine learning approach that combines multiple classification algorithms to achieve better performance than could be achieved by any of the constituent algorithms alone. It has been reported for obtaining maximum predictive performance and has been applied in many areas of medical diagnosis, from neurodegenerative diseases (25, 26) and cancer (27–29) to regenerative medicine for identifying stem cell differentiation (30). Here, we supplied our SCRS to eight different classification models, namely LDA, XGB (extreme gradient boosting), SVM-Linear (support vector machine with a linear kernel), SVM-Radial (support vector machine with a radial basis function kernel), MLPNN (monotune multi-layer perceptron neural network), RF (random forest), MDA (mixture discriminant analysis) and GBM (gradient boosting machine). Then, a separate GBM was employed to ensemble the outputs from the different classifiers and establish an ensemble learner which gave the final diagnosis (Figure 4). The eight individual classifiers were found to have low correlations with each other (Figure S2); therefore, the ensemble learner could take full advantage of different classifiers without repetitive computational costs.

**Figure 4.**
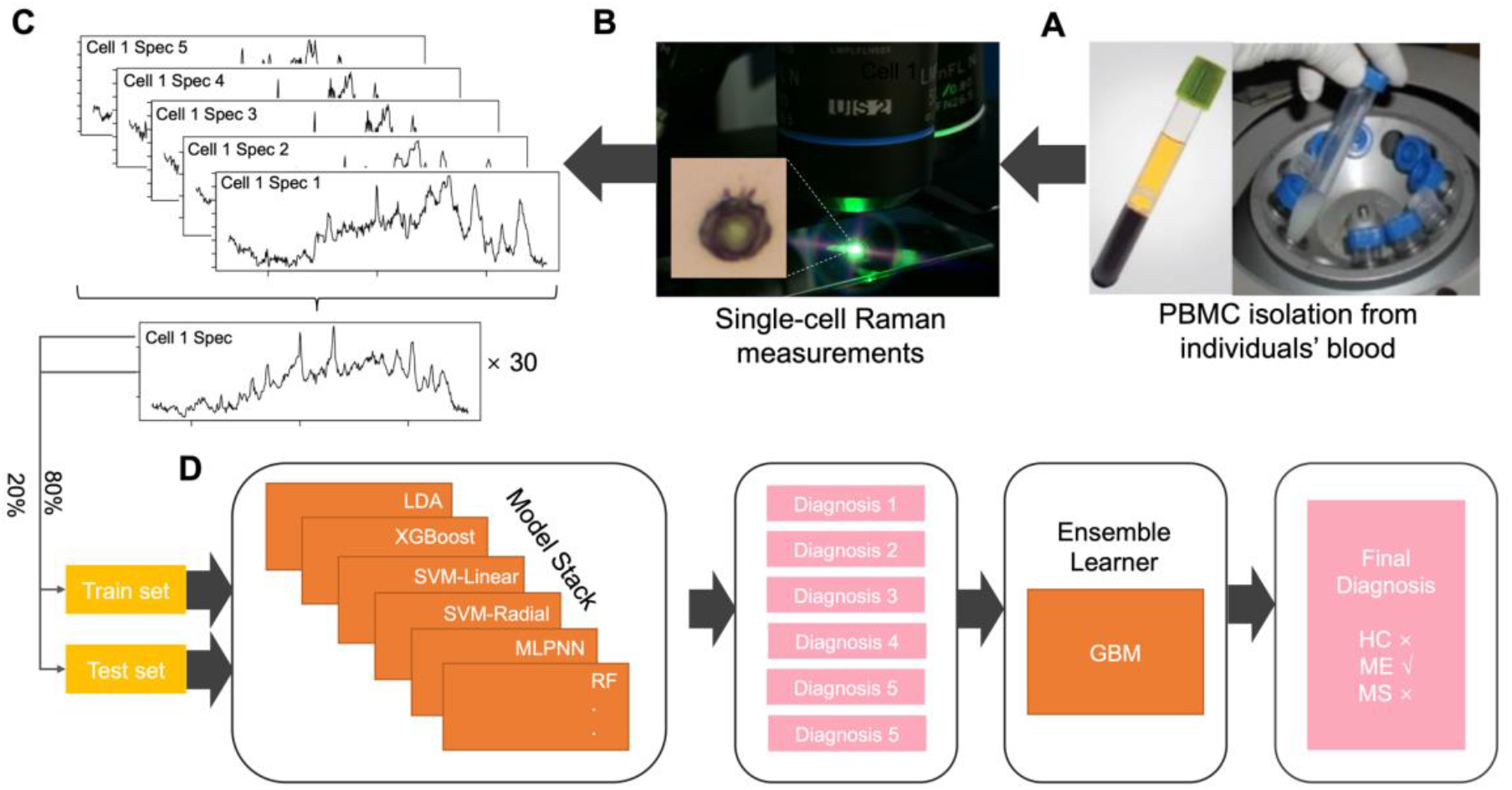
Schematic illustration of the blood-based Raman spectroscopic diagnostic test for ME/CFS and MS at a single-cell level. **(A)** PBMCs were isolated from blood samples. **(B)** Raman spectra of single PBMCs from 98 individuals were measured. **(C)** Around 5–7 spectra were measured in each cell which were then averaged to one spectrum for one cell. **(D)** SCRS from 98 individuals were then split into a train set (80%) and a test set (20%) with balanced subgroup distribution. The train set was used to train an ensemble learner and the independent test set was input into the trained learner for diagnosing the cell as HC, ME or MS.

Figure 4 shows the technical, spectroscopic and machine learning workflow of a blood cell-based diagnostic test using SCRS of PBMCs and an ensemble learning machine learning model. After the isolation of PBMCs from the patient’s blood, cells are taken for Raman spectroscopic examination. Single-cell spectra are averaged from >5 spectra per cell and put into the trained ensemble learning classification model for individual diagnoses. The individual diagnoses are then further converted via a GBM model to give a final diagnosis.

The multi-model ensemble learner showed effectively much better performance for the five-class prediction task on the independent test set (Figure 5A). The accuracy was significantly improved from 47–61% in individual models (Table S5) to 83.8% in the ensemble model (Figure 5A), demonstrating the enhanced predictive power of the ensemble model by combining different learning algorithms. Specifically, the ensemble model showed capacity in differentiating subgroups of ME/CFS patients based on their symptom severity. The sensitivity and specificity are 88% and 95% for the mild, 86% and 98% for the moderate, and 71% and 97% for the severe (Figure 5A). Around 12% of the severe ME patients have been classified as either moderate or mild, while 11% have been classified as MS, suggesting a resemblance of the two diseases, especially MS and ME/CFS with high severity (Figure 5A).

**Figure 5.**
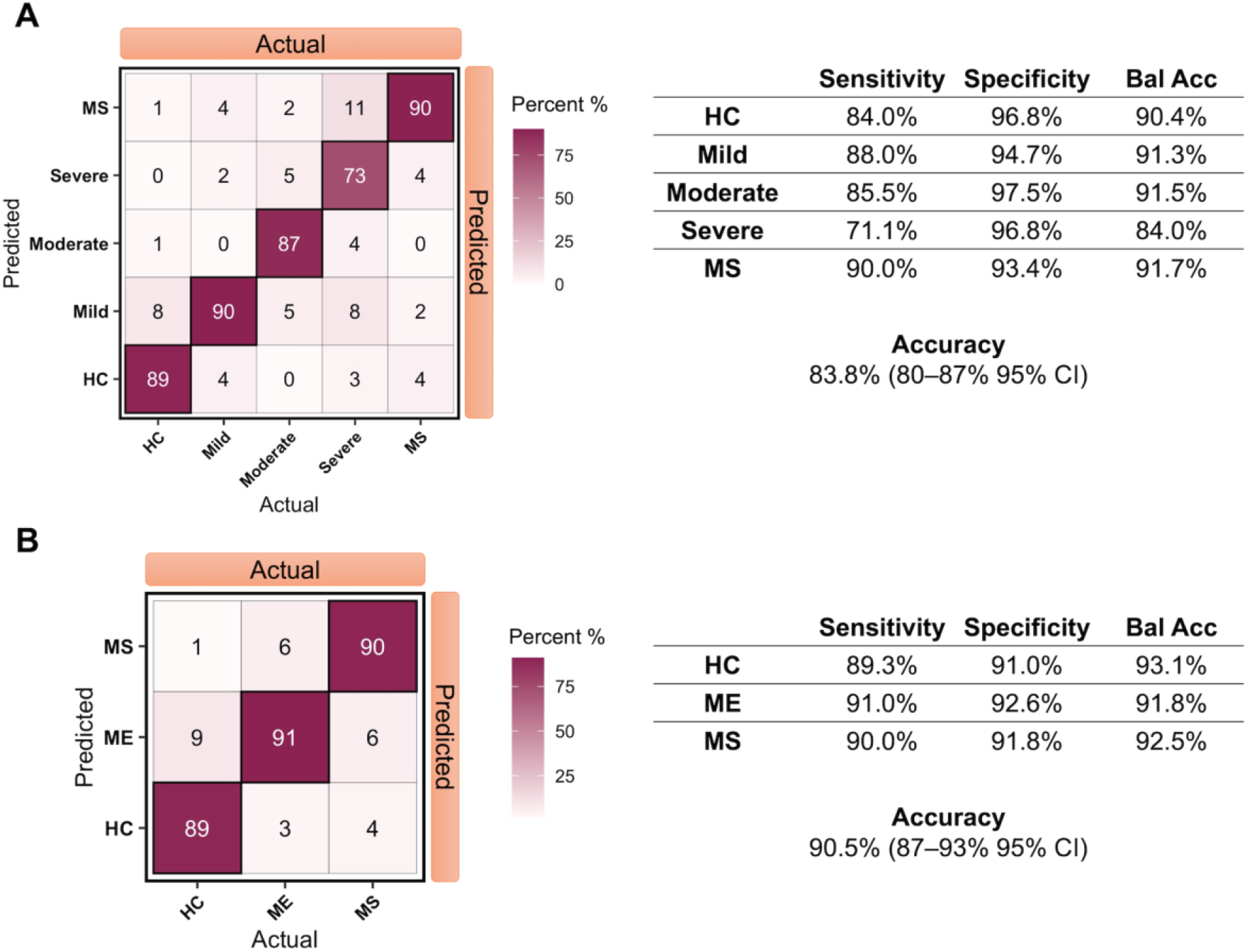
Ensemble learner performance on an independent test set breakdown by **(A)** five classes with 84% overall accuracy and **(B)** three classes with overall 91% accuracy. Matrix entries are shown as percentage values. The three-class classification model shows a performance of diagnosing ME/CFS with 91% sensitivity and 93% specificity, MS with 90% sensitivity and 92% specificity, and an overall accuracy at 91% with 87–93% at 95% confidence interval.

For better clinical relevance on the predictive perspective, we also looked at the performance of the ensemble learner for the three-class classification tasks, that is, classifying each single-cell spectrum as either MS, ME/CFS or HC (Figure 5B). The model achieved high performance on the independent test set with a sensitivity of 91% and specificity of 93% for the ME/CFS group; a 90% sensitivity and 92% specificity for the MS group. The overall accuracy on the test set was 91% (87–93% at a 95% confidence interval) (Figure 5B). These results support our approach of a blood cell-based diagnostic test combining single-cell Raman spectroscopy and ensemble learning as a diagnostic tool for ME/CFS.

## Discussion

In this work, we acquired SCRS of PBMCs from 98 individuals’ blood for a diagnostic test of ME/CFS and MS. A sample-to-diagnosis workflow was established, which involves PBMC isolation, Raman spectroscopic measurements, ensemble learning classification, and final diagnosis. The SCRS of PBMCs from healthy controls, ME/CFS and MS disease controls showed distinct characteristics, and different phenotypes were visualized as clear clusters at both single-cell level and individual level. None of the FSS scale, clinical symptom presentations, nor assessment of mitochondrial respiration could separate the different severity classes of ME/CFS patients from MS patients and healthy controls, suggesting the need for a simple and direct diagnostic test for the disease. An SF-36 score of 25 was used to separate the mild and moderate groups with good separation by Raman microscopy. Future studies will explore the use of MFI-20 and DSQ questionnaires which could potentially improve the mild and moderate diagnoses. However, patients can fluctuate between mild and moderate states, hence, assigning patients into one of the two groups will be a challenge.

By quantifying levels of intracellular metabolites via PBMCs’ Raman spectra, we observed metabolic differences the ME/CFS and two control cohorts. Most of the metabolic changes that have been reported in previous ME/CFS studies of plasma are consistent with direct and indirect effects of energy strain (31–35) and abnormal lipid metabolism (34–38). Our findings agree with the altered utilization of amino acids in patients with ME/CFS, including AAAs of tryptophan, tyrosine, and phenylalanine. This was also shown in the MS group compared to healthy controls. Tryptophan, a necessary amino acid that causes significant changes in mood and fatigue, is the precursor to serotonin and kynurenine. There was a particular decrease in the tryptophan in all disease groups, possibly suggesting changes in the kynurenine pathway and NAD biosynthesis (32). Reduced neuroactive tryptophan metabolites could induce central fatigue via neuronal mechanisms, which is a hallmark of both ME/CFS and MS (39). It has been proposed that high levels of tryptophan in the immune cells of ME/CFS patients could link to a metabolic trap hypothesis (40). The hypothesis proposes that patients are unable to generate kynurenine due to mutations in the Indoleamine 2,3-Dioxygenase 2 (IDO2) gene, causing a build-up of tryptophan inhibiting the production of kynurenine, via repression of the more catalytically active IDO1 isozyme. Our data also suggests a build-up of tryptophan in the PBMC cell fractions in ME/CFS patients. However, PBMC fractions contain mixed cell populations; cells expressing IDO1 and IDO2, including myeloid and plasmacytoid dendritic cells, only make up a small percentage of the PBMC fraction. In the future, this study should be continued using IDO1 and IDO2-expressing cell types.

Together with the observation of decreased tyrosine levels in all disease groups and reduced phenylalanine in the severe ME group, our findings are consistent with other studies of patient metabolism using broad-spectrum metabolite analysis which indicated abnormal metabolite levels in patients with reduced levels of serum amino acids and disturbances in pyruvate dehydrogenase, sphingolipid and phospholipid metabolism (13, 33, 34, 41). Elevated glycerol was observed in all subgroups of ME and the MS group. In combination with the altered unsaturated fatty acid levels in the ME and MS groups, this may suggest that lipolysis is induced where glycerol is broken down into free fatty acids, usually occurring during times of energy deprivation, such as fasting and exercising (42, 43). Altered utilisation of fatty acids and amino acids also suggests an alternative strategy for energy fuelling. Glycogen and glucose levels decreased in cells of all disease groups. A well-known mechanism of fatigue is the progressive rise in muscle amino acid oxidation rates that occurs after continuous exercise, which leads to glycogen depletion (44, 45). Altered fatty acid metabolism and reduced cholesterol and cholesteryl esters were observed. There has been evidence indicating that changes in lipid metabolism, such as those in cholesterol, sphingolipids, and fatty acids, have a role not only in the aetiology of neurodegenerative diseases like MS but also as indicators of the disease’s occurrence and development (46). Unusual lipid-mediated communication in immune cells is one theory that could explain the aetiology of MS (47).

We explored the use of ensemble learning for the identification of cells based on their Raman spectra. The ensemble learner achieved good predictive performance when tested via a sound train/test split protocol and was improved significantly by combining individual learning algorithms. It achieved an accuracy rate of 91% for making a diagnosis of either MS, ME/CFS or non-fatigue; and 84% accuracy with additionally identifying ME disease severeness. The ensemble learner pre-training method can be easily extended to new clinical settings for diagnosis. As the test was carried out at the single-cell level, only a small amount of blood sample is required. It also has the benefit of being able to be carried out on fixed material without the limitations of a live cell assay. A simple and minimally invasive test by analysis of blood cells for the diagnosis of fatigue syndromes like ME/CFS in primary care has the potential to make drastic impacts on patients’ quality of life. Of particular interest is the achievement of 91% sensitivity and 93% specificity for ME/CFS group while, to date, diagnosis and assessment of ME/CFS in research studies are often based upon self-report, questionnaires and subjective measures, e.g., fatigue severity and impact (48). An objective, sensitive and straightforward diagnostic tool can therefore resolve the controversy concerning the nature of ME/CFS and make a significant difference in the medical, economic, social and emotional impact of ME/CFS on individual patients and society. Although single-cell Raman spectroscopic approach is not yet readily available in certified diagnostic laboratories, which might represent a barrier to adoption, our study investigates its potential as a brand-new diagnostic technique that is rapid (measurement for one sample < 1 h), accurate (91% accuracy) and minimally invasive thus allowing for more often longitudinal monitoring of the diseases. Validation of this approach involving more testing subjects and further optimisations will follow this study in an attempt to provide additional evidence of its diagnostic potential in generalised clinical settings.

Cryopreservation has previously been shown to have impacts on immune cell markers (49, 50) as well as functions (51, 52). Contrary to the impact of cryopreservation on PBMC functionality, the application of single-cell Raman micro-spectroscopy using frozen samples, fixed prior to analysis, was extremely robust, giving the excellent differentiation of the patient groups with the additional ability to separate the different ME/CFS severity types. Compared with other blood tests being explored in ME/CFS (53–55), our Raman approach on fixed PBMC samples has many benefits. Once prepared, samples can be stored in liquid nitrogen or at −80°C for a prolonged time frame and the freezing duration did not show a significant effect contributing to classification using the Raman dataset. Future studies should also compare freshly fixed PBMC compared to samples cryopreserved before fixation. This was not possible in this study due to samples being obtained frozen from the ME/CFS UK biobank. Examining freshly fixed samples in the future would be a better assessment of how this approach might be used clinically as a diagnostic test.

Examining mitochondrial or energetic functions of PBMC has shown promise as a test for PBMC. Still, results have varied between labs with the significant problems encountered when developing a live cell test, such as low energetic activities of the cells. The ability to use fixed PBMCs is a major benefit allowing samples to be stored easily prior to analysis. The single-cell nature of the Raman spectroscopic approach only requires a small blood sample which could be developed as a point-of-care test perhaps from one drop of blood. If differences can be identified in plasma or serum, this approach would be provided with a sample which is easy to generate, and a much faster output could be given.

Over the next decade, advances in healthcare technologies and a greater focus on prevention, early diagnosis and well-being will bring major improvements in patient outcomes. In chronic unexplained conditions such as ME/CFS, using complex data and machine learning approaches will be of great importance not only in diagnosis but also in defining the early stages of the conditions where the patient shows no or few symptoms. This will help in establishing new treatment approaches linked to subtle changes in the patients’ biology, preventing the development of the full-blown disease, which is harder to treat once established. Predictive analytics based on Auto prognosis, a tool generalising risk scores using machine learning algorithms, have already been tested in a number of clinical settings (56). To our knowledge, together with our pilot study, this is the first research using Raman spectroscopy and advanced machine learning techniques to discriminate subgroups of ME/CFS patients based on the symptoms severity, achieved with high accuracy, sensitivity, and specificity. With sophisticated machine learning algorithms, our Raman spectroscopic approach has great potential as a diagnostic technique for diseases like ME/CFS. With ongoing plans of expanding our cohort size and bringing in other cohorts of patients with similar co-morbidities, Raman spectroscopy coupled with other techniques, such as metabolomics and proteomics, can further expand our knowledge on the diseases and potentially identify common disease pathways or drivers.

## Methods

### Ethics

This study was approved by the Research Ethics Committees at the University of Oxford (Reference number: R51826/RE001) and by the UCL Biobank Ethical Review Committee-Royal Free (B-ERC-RF) London NHS Foundation Trust (Reference number: EC.2017.012). Ethical approval for sample and data collection and storage was granted by the London School of Hygiene & Tropical Medicine (LSHTM) Ethics Committee (Ref. 6123) and the National Research Ethics Service (NRES) London-Bloomsbury Research Ethics Committee (REC ref. 11/10/1760, IRAS ID:77765). Samples were provided by the UK ME/CFS Biobank (UKMEB) in accordance with a Material Transfer Agreement signed by the London School of Hygiene & Tropical Medicine and the University of Oxford.

### Statistics and clinical measurements

Descriptive statistics (median and range) were calculated to summarise sociodemographic variables. All continuous clinical variables were analysed using two-sided non-parametric tests for independent samples; Wilcoxon rank sum test with continuity correction (Mann Whitney U), with categorical variables compared using Fisher’s Exact Test.

Symptom presence and intensity was determined for 63 variables obtained from the UKMEB symptoms assessment. Responses were recorded on an ordinal 4-point scale, for all three groups (HC, MS, ME), with 0 indicating “absent”, 1 indicating “mild”, 2 indicating “moderate” and 3 indicating “severe”. Variables were worded such as “Have you had dizziness standing up?”, or “Have you had confusion/brain fog?”. Clinical data was analysed using R version 4.0.2. Heatmaps were created using the ggplot2 package.

### PBMC processing

PBMC were obtained from the UK ME/CFS Biobank London School of Hygiene & Tropical Medicine and stored at −80°C until needed. The PBMC samples used were processed according to the UCL-RFH Biobank protocol (BB/SOP/007/01) by the personnel at the UCL-RFH Biobank laboratory.

The process includes dilution with sterile HBSS, separation on centrifugation using Lympholyte as a medium, and the isolation of mononuclear cells which are pelleted following further centrifugation. Samples had been stored between 1 and 5 years at the Biobank in liquid nitrogen tanks.

### Oxygen consumption rate (OCR) assays

Cells were gently de-frosted and counted using Trypan Blue. For the oxygen consumption rate (OCR) assay, 500,000 cells per well were plated in 96-well black plates. MitoXpress Xtra Oxygen sensing probe (Agilent) was added according to the manufacturer’s instructions, using warmed High Sensitivity (HS) mineral oil. The plates were read by ClarioStar plate reader (BMG Labtech) in time-resolved fluorescence mode using Ex TR-ex filter (340 nm) and Em TR-em (650 nm), at 37 °C. OCR was calculated using MARS Software (BMG Labtech). For Raman micro-spectroscopic measurements, 5,000 cells were washed in phosphate buffered saline (PBS) and fixed in 4% paraformaldehyde (PFA) for 15 minutes then washed with PBS, centrifuged and pellets re-suspended in PBS, stored at 4 °C until analysed.

### Measurements of single-cell Raman spectra (SCRS)

Raman spectroscopic measurements were blinded in this study. Upon receival, the PFA-fixed cells were washed twice with dH_2_O to remove traces of medium or chemical. Then 2 μl of each sample was dropped onto an aluminium-coated microscopic slide to be air-dried. Raman spectra were acquired using an HR Evolution confocal Raman microscope (Horiba Jobin–Yvon, UK Ltd) equipped with a 532 nm neodymium–yttrium aluminum garnet laser. The laser power on the cells was 5 mW, attenuated using neutral density filters. An air objective (50×, NA = 0.65) was used to focus single cells. Raman scattering from the focal point was detected by a charge coupled device (CCD) cooled at −70 °C. SCRS were measured from 320 to 3400 cm^−1^ with a 300 grooves/mm grating. A mapping mode was used to characterise single cells and the acquisition parameters were 5 s per spectrum. As each cell sized around 3–10 μm^2^ after air-dried, 5–10 spectra per cell and ∼30 single cells per each group were measured. After quality control to remove spectra with low signal-to-noise ratios, measurements of 98 samples yielded a total number of 14600 Raman spectra from 2155 single cells. The spectra from each cell were than averaged into SCRS.

### Preprocessing of SCRS

All spectra were preprocessed by comic ray correction, polyline baseline fitting and vector normalisation of the entire spectral region. Data analysis, statistics and visualisation were done in an R environment (4.0.0). Quantification of intracellular biomolecules was done by integrating the corresponding Raman bands in SCRS. Raman bands were integrated at 758, 880, 1013 and 1550 cm^−1^ and 1022–1036 cm^−1^ for quantifying tryptophan; 1003 and 1032 cm^−1^ for quantifying phenylalanine; 642, 830 and 850 cm^−1^ for quantifying tyrosine. The quantification results were represented as box plots and sample means were compared with healthy control (HC) by using Welch’s two sample t-test for unequal variance.

### Ensemble learning classification model

SCRS were divided into a train set and a test set with a 0.8:0.2 split ratio. Both the train and test sets contained a balanced number of samples from five groups of MS, Severe ME, Moderate ME, Mild ME and HC. While the train set was used to train a classification model, the test set was used to independently evaluate the model performance. A cross-validation approach was used to enable all samples to enter the independent test set at least once, therefore, making the best use of the sample pool. The final performance measurements were reported based on averages on all cross-validation results. Pre-processing of the raw Raman spectra involved scaling, centring and dimension reduction by principal component analysis to the first hundred principal components. The pre-processed Raman spectra were used as the inputs into eight different classification models, namely LDA, XGB (extreme gradient boosting), SVM-Linear (support vector machine with linear kernel), SVM-Radial (support vector machine with radial basis function kernel), MLPNN (monotune multi-layer perceptron neural network), RF (random forest), MDA (mixture discriminant analysis) and GBM (gradient boosting machine). Tenfold cross-validation with five repetitions was used during model construction. After establishing and evaluating the classification models based on the eight classifiers, eight models were stacked together to build a two-layer ensemble learner for a better classification result. The prediction outcomes of the test set from the eight models were used as features for the ensemble learner which used a GBM. All models were constructed in an R 4.0.0 environment.

## Data Availability

All data produced are available online at

https://doi.org/10.6084/m9.figshare.14892051.v1.

## Acknowledgements

This research project was support by an ME Association project grant to KM. We thank Professor Antonyia Georgieva and Dr Jonathan Williams for critically reviewing this manuscript.

## Author Contributions

K.J.M. and W.E.H. contributed conceptualization; J.X. and K.J.M. wrote original draft; all authors contributed to reviewing and editing of the final manuscript. J.X. performed Raman data acquisition, analysis and machine learning models. T.L. carried out the mitochondrial experiments and processing samples. C.K. and E.L. recruited patients, processed blood samples and supported the clinical analysis. J.S. carried out the clinical statistics and modelling. J.M. supported the MS component of the study incorporating into the wider study. P.Z. supported the interpretation of the clinical assessments.

## Data Availability

Data used in this study is available at https://doi.org/10.6084/m9.figshare.14892051.v1.

## Supporting Information

Figure S1 and S2, Tables S1, S2, S3, S4 and S5 are included in Supporting Information.

