## Supporting Information for "Developing a blood cell-based diagnostic test for myalgic encephalomyelitis/chronic fatigue syndrome using peripheral blood mononuclear cells"

**This PDF file includes:**

Figures S1 to S2

Tables S1 to S5

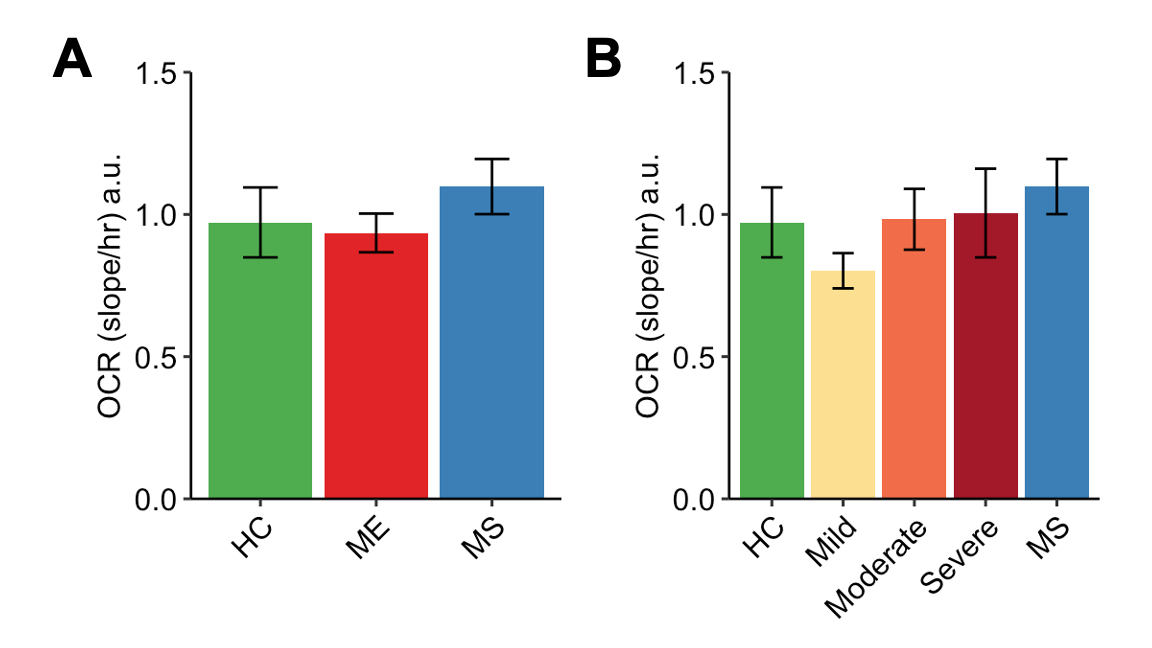

**Fig. S1**. OCR of PBMCs from 41 human subjects of **(A)** HCs (n = 9), ME patients (n = 26) and MS (n = 6) patients, and **(B)** HCs and MS patients, with ME patients separated based on disease severities of Mild (n = 9), Moderate (n = 8), and Severe (n = 9). No significant statistical difference between HCs and patients were observed.

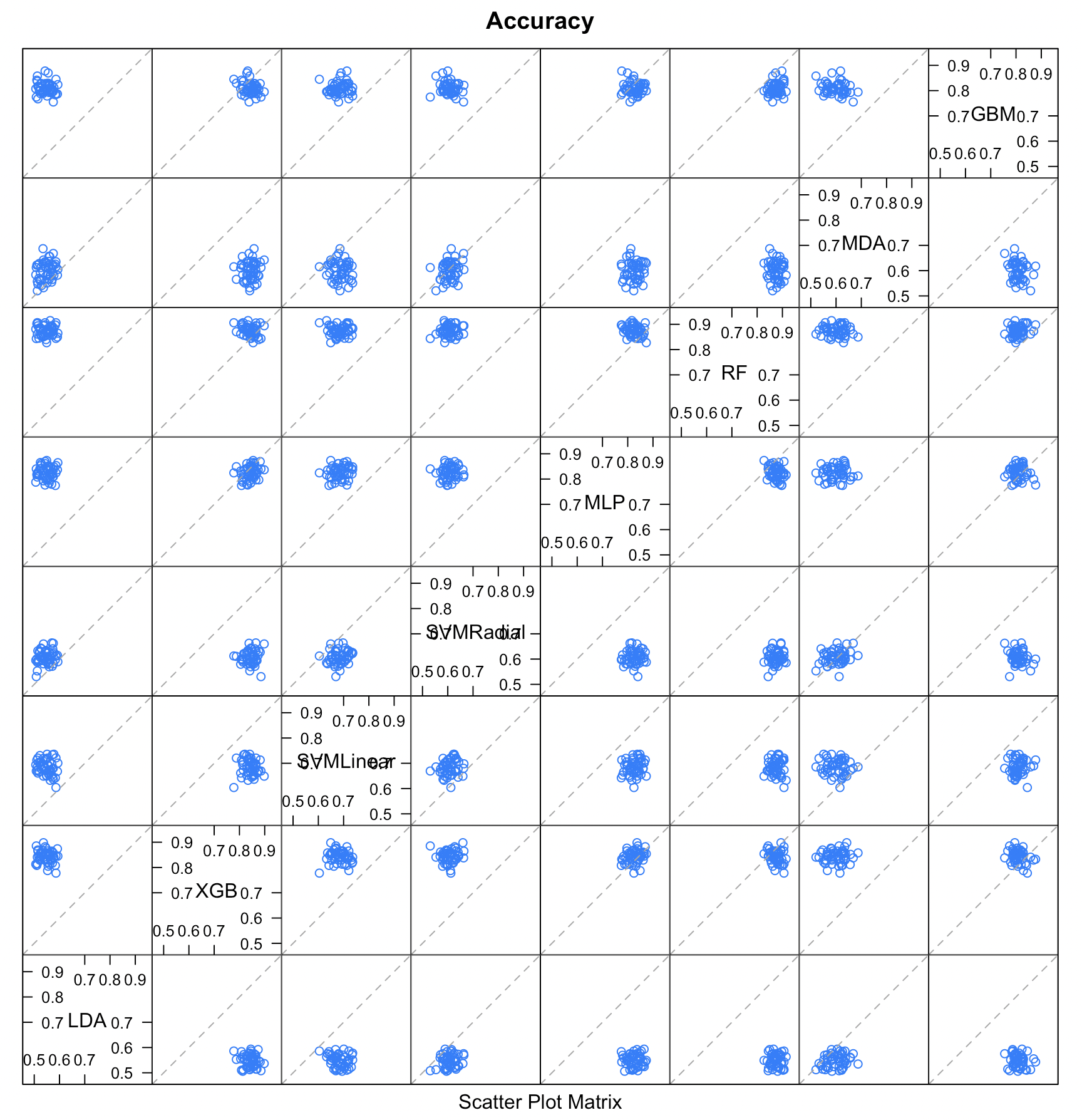

**Fig. S2.** Model correlations by comparing model accuracies. LDA: Linear Discriminant Analysis; kNN: k-Nearest Neighbour; SVM-Linear: Support Vector Machine with Linear Kernel: SVM-Radial: Support Vector Machine with Radial Basis Function Kernel; MLPNN: Monotone Multi-Layer Perceptron Neural Network; RF: Random Forest; MDA: Mixture Discriminant Analysis; XGB: Extreme Gradient Boosting; GBM: Stochastic Gradient Boosting

**Table S1**. Retrospective subject cohort breakdown with fatigue groups in this study. Gender: F represents Female and M represents Male; Fatigue Severity Scale (FSS), General Health Questionnaire 28 (GHQ-28). Statistical comparison was shown in the blue area. Comparisons of 2 groups was performed using the Mann Whitney U test (Wilcoxon rank sum test with continuity correction). Comparisons of greater than 2 groups employed the non-parametric Kruskal Wallis rank sum test. Fischer's exact test was used to compare categorical variables. Comparisons with statistical significance where p < 0.05 were highlighted.

| **Variable** | **Gender (M/F)** | **Age** | **Disease Duration (years)** | **Body Mass Index** | **FSS** | **Somatic Symptoms(GHQ-28:1-7)** | **Anxiety & Insomnia(GHQ-28:8-14)** | **Social Dysfunction(GHQ-28:15-21)** | **Severe depression(GHQ-28:22-28)** | **GHQ-28:1-28 (Sum)** |
| --- | --- | --- | --- | --- | --- | --- | --- | --- | --- | --- |
| **ME (Mild) Median** | 4/20 | 46 | 12.4 | 25.5 | 57 | 2.5 | 1.5 | 1 | 0 | 4 |
| Range | 0-1 | 18-59 | 0.42-37.9 | 17.9-46.7 | 44-63 | 0-7 | 0-6 | 0-6 | 0-7 | 0-26 |
| **ME (Moderate) Median** | 1/11 | 45.5 | 9 | 32.4 | 62.5 | 3 | 2 | 4 | 0 | 8 |
| Range | 0-1 | 30-60 | 1.58-29.2 | 25.1-53.8 | 44-63 | 0-5 | 0-5 | 0-7 | 0-6 | 0-17 |
| **ME (Severe) Median** | 6/14 | 40 | 15.1 | 21.5 | 59.5 | 2.5 | 1 | 1.5 | 0 | 6.5 |
| Range | 0-1 | 23-59 | 1.5-39.8 | 17.2-32.2 | 54-63 | 0-7 | 0-6 | 0-7 | 0-5 | 0-23 |
| **ME (All/Pooled) Median** | 11/45 | 42.5 | 12.4 | 26.2 | 59 | 3 | 1 | 2 | 0 | 7 |
| Range | 0-1 | 18-60 | 0.42-39.8 | 17.2-53.9 | 44-63 | 0-7 | 0-6 | 0-7 | 0-7 | 0-26 |
| **Multiple Sclerosis Median** | 6/16 | 51.5 | 15.5 | 23.8 | 54 | 3 | 1 | 3 | 0 | 9 |
| Range | 0-1 | 38-60 | 1.17-39.2 | 20.7-37.7 | 16-63 | 0-7 | 0-7 | 0-7 | 0-6 | 0-25 |
| **Healthy Controls Median** | 6/12 | 43 | / | 26.6 | 17 | 0 | 0 | 0 | 0 | 0 |
| Range | 0-1 | 18-60 | / | 20.2-36.4 | 11-37 | 0-3 | 0-3 | 0-4 | 0-1 | 0-8 |
| **Across ME Severities (p value)** | 0.36 | 0.7601 | 0.5446 | **0.0028** | **0.0667** | 0.5764 | 0.6049 | 0.2482 | 0.9371 | 0.7765 |
| **ME (All severities) vs MS (p value)** | 0.55 | **0.0098** | 0.2898 | 0.3141 | **0.0017** | 0.53 | 0.8275 | 0.2532 | 0.6404 | 0.3546 |
| **Healthy Controls vs ME (All severities) p value** | 0.3331 | 1 |  | 0.9949 | **0** | 0.0022 | 9e-04 | 0.0119 | **0.0287** | 5e-04 |
| **Healthy Controls vs Multiple Sclerosis (MS) p value** | 0.7385 | **0.0238** |  | 0.2793 | **0** | 9e-04 | 0.0036 | 0.0014 | **0.0225** | 0 |

**Table S2**. UKMEB symptom burden assessment. Symptom feature inclusion was determined by calculating the relative mean ordinal intensity for each variable to provide sufficient detail to rank order (as opposed to a median derived integer with categorical representation) symptoms, allowing for selective inclusion with a >1.5-fold difference between groups (severe ME compared to MS). Air hunger: adjusted for confounding administration of Asthma medications salbutamol and or a steroidal inhaler (n=7 subjects across experimental groups): 0.000251253 (p), 0.008040096 (p) (Hochberg adjusted for multiple comparisons).

| **UKMEB Full Variable** | **UKMEB Short Name** | **Relative ordinal score (Severe ME compared to MS)** | **Mean ordinal score (HCs)** | **Range ordinal score (HCs)** | **NA Count total (%) (HCs)** | **Mean ordinal score (MS)** | **Range ordinal score (MS)** | **NA Count total (%) (MS)** | **Mean ordinal score (Severe ME)** | **Range ordinal score (Severe ME)** | **NA Count total (%) (Severe ME)** | **p-value (Fisher's Exact Test)** | **p-value (Hochberg adjusted)** |
| --- | --- | --- | --- | --- | --- | --- | --- | --- | --- | --- | --- | --- | --- |
| PQ4.9.42 Over past week, have you had worsening symptoms after exertion? | Symptoms worsening after exertion | 2.16 | 0.06 | 0 - 1 | 1 (5.56) | 1.32 | 0 - 3 | 0 (0) | 2.85 | 2 - 3 | 0 (0) | **0.000001** | **0.000019** |
| PQ4.9.8 Over past week, have you felt ill (>24hrs) after exertion? | Feeling ill (>24hrs) after exertion | 2.59 | 0.00 | 0 - 0 | 1 (5.56) | 0.77 | 0 - 2 | 0 (0) | 2.00 | 2 - 2 | 0 (0) | **0.000004** | **0.000112** |
| PQ4.9.41 Over past week, have you had exercise intolerance? | Exercise intolerance | 1.77 | 0.06 | 0 - 1 | 1 (5.56) | 1.64 | 0 - 3 | 0 (0) | 2.90 | 2 - 3 | 0 (0) | **0.000013** | **0.000400** |
| PQ4.9.9 Over past week, have you had pain (>24hrs) after exertion? | Pain (>24hrs) after exertion | 2.89 | 0.06 | 0 - 1 | 1 (5.56) | 0.76 | 0 - 2 | 1 (4.55) | 2.20 | 0 - 3 | 0 (0) | **0.000023** | **0.000653** |
| PQ4.9.6 Over past week, have you had new sensitivities? | New sensitivities | 4.03 | 0.06 | 0 - 1 | 1 (5.56) | 0.55 | 0 - 3 | 0 (0) | 2.20 | 0 - 3 | 0 (0) | **0.000042** | **0.001181** |
| PQ4.9.19 Over past week, have you had air hunger? | Air hunger | 3.80 | 0.00 | 0 - 0 | 1 (5.56) | 0.50 | 0 - 3 | 0 (0) | 1.90 | 0 - 3 | 0 (0) | **0.000079** | **0.002133** |
| PQ4.9.4 Over past week, have you had tender glands in neck/arm pit? | Tender glands in neck/arm pit | 4.95 | 0.06 | 0 - 1 | 2 (11.11) | 0.33 | 0 - 3 | 1 (4.55) | 1.65 | 0 - 3 | 0 (0) | **0.000106** | **0.002753** |
| PQ4.9.39 Over past week, have you had fatigue (>24hr) after exertion? | Fatigue (>24hrs) after exertion | 1.52 | 0.00 | 0 - 0 | 1 (5.56) | 1.32 | 0 - 2 | 0 (0) | 2.00 | 2 - 2 | 0 (0) | **0.000204** | **0.005101** |
| PQ4.9.21 Over past week, have you had new headaches? | New headaches | 2.05 | 0.00 | 0 - 0 | 1 (5.56) | 1.00 | 0 - 3 | 0 (0) | 2.05 | 0 - 3 | 1 (5) | **0.000261** | **0.006258** |
| PQ4.7b In past week, how likely are you to doze off while watching tv? | Likelihood of dozing off while watching TV | 0.34 | 1.24 | 0 - 3 | 1 (5.56) | 2.18 | 0 - 3 | 0 (0) | 0.75 | 0 - 3 | 0 (0) | **0.000494** | **0.011367** |
| PQ4.9.35 Over past week, have you had slow thinking? | Slow thinking | 1.56 | 0.18 | 0 - 1 | 1 (5.56) | 1.45 | 0 - 3 | 0 (0) | 2.26 | 2 - 3 | 1 (5) | **0.000711** | **0.015641** |
| PQ4.9.2 Over past week, have you had flu symptoms? | Had flu symptoms | 4.63 | 0.29 | 0 - 2 | 1 (5.56) | 0.27 | 0 - 3 | 0 (0) | 1.26 | 0 - 3 | 1 (5) | **0.002296** | **0.048215** |
| PQ4.9.23 Over past week, have you had sensitivity to light/noise/smell/touch? | Sensitivity to noise/smell/touch | 2.31 | 0.12 | 0 - 2 | 1 (5.56) | 0.91 | 0 - 3 | 0 (0) | 2.10 | 0 - 3 | 0 (0) | **0.002787** | 0.055738 |
| PQ4.9.37 Over past week, have you had unrefreshing sleep? | Unrefreshing sleep | 1.67 | 0.41 | 0 - 1 | 1 (5.56) | 1.50 | 0 - 3 | 0 (0) | 2.50 | 1 - 3 | 0 (0) | **0.011295** | 0.205433 |
| PQ4.9.10 Over past week, have you had muscle pain? | Muscle pain | 1.56 | 0.47 | 0 - 2 | 1 (5.56) | 1.38 | 0 - 3 | 1 (4.55) | 2.16 | 0 - 3 | 1 (5) | **0.011413** | 0.205433 |
| PQ4.9.28 Over past week, have you had short term memory problems? | Short term memory problems | 1.55 | 0.29 | 0 - 2 | 1 (5.56) | 1.45 | 0 - 3 | 0 (0) | 2.25 | 1 - 3 | 0 (0) | **0.013710** | 0.233064 |
| PQ4.9.1 Over past week, have you had a sore throat? | Sore throat | 2.57 | 0.35 | 0 - 3 | 1 (5.56) | 0.41 | 0 - 3 | 0 (0) | 1.05 | 0 - 3 | 1 (5) | **0.017284** | 0.233719 |
| PQ4.7c In past week, how likely are you to doze off while sitting inactive? | Likelihood of dozing off while sitting inactive | 0.31 | 0.24 | 0 - 1 | 1 (5.56) | 1.59 | 0 - 3 | 0 (0) | 0.50 | 0 - 3 | 0 (0) | **0.018914** | 0.233719 |
| PQ4.9.47 Over past week, have you had palpitations other times? | Palpitations at times other than while standing | 2.97 | 0.09 | 0 - 1 | 7 (38.89) | 0.45 | 0 - 2 | 0 (0) | 1.35 | 0 - 3 | 0 (0) | **0.025242** | 0.233719 |
| PQ4.9.50 Over past week, have you been unusually sweaty? | Been unusually sweaty | 3.54 | 0.24 | 0 - 1 | 1 (5.56) | 0.41 | 0 - 3 | 0 (0) | 1.45 | 0 - 3 | 0 (0) | **0.025886** | 0.233719 |
| PQ4.9.7 Over past week, have you had alcohol intolerance? | Alcohol intolerance | 2.56 | 0.08 | 0 - 1 | 6 (33.33) | 0.64 | 0 - 3 | 0 (0) | 1.63 | 0 - 3 | 1 (5) | **0.041215** | 0.233719 |
| PQ4.9.46 Over past week, have you had palpitations while standing? | Palpitations while standing | 3.46 | 0.06 | 0 - 1 | 1 (5.56) | 0.32 | 0 - 2 | 0 (0) | 1.10 | 0 - 3 | 0 (0) | **0.043068** | 0.233719 |
| PQ4.9.15 Over past week, have you had pain in >=2 joints? | Pain in 2 or more joints | 1.65 | 0.29 | 0 - 2 | 1 (5.56) | 1.09 | 0 - 3 | 0 (0) | 1.80 | 0 - 3 | 0 (0) | **0.044137** | 0.233719 |
| PQ4.9.31 Over past week, have you had disorientation? | Disorientation | 2.04 | 0.18 | 0 - 2 | 1 (5.56) | 0.77 | 0 - 2 | 0 (0) | 1.58 | 0 - 3 | 1 (5) | 0.053621 | 0.233719 |
| PQ4.9.14 Over past week, have you had pain in chest/abdomen? | Pain in chest/abdomen | 1.95 | 0.08 | 0 - 1 | 6 (33.33) | 0.73 | 0 - 3 | 0 (0) | 1.42 | 0 - 3 | 1 (5) | 0.080233 | 0.233719 |
| PQ4.9.45 Over past week, have you had dizziness while standing? | Dizziness while standing | 1.75 | 0.12 | 0 - 1 | 1 (5.56) | 1.00 | 0 - 3 | 0 (0) | 1.75 | 0 - 3 | 0 (0) | 0.084262 | 0.233719 |
| PQ4.9.48 Over past week, have you felt light-headed? | Felt light-headed | 1.98 | 0.18 | 0 - 1 | 1 (5.56) | 0.86 | 0 - 3 | 1 (4.55) | 1.70 | 0 - 3 | 0 (0) | 0.091235 | 0.233719 |
| PQ4.9.49 Over past week, have you been extremely pale? | Been extremely pale | 2.20 | 0.12 | 0 - 2 | 1 (5.56) | 0.73 | 0 - 3 | 0 (0) | 1.60 | 0 - 3 | 0 (0) | 0.096796 | 0.233719 |
| PQ4.9.3 Over past week, have you had fever/chills? | Had fever/chills | 2.89 | 0.18 | 0 - 2 | 1 (5.56) | 0.45 | 0 - 3 | 0 (0) | 1.32 | 0 - 3 | 1 (5) | 0.097901 | 0.233719 |
| PQ4.9.5 Over past week, have you had any viral infections? | Any viral infections | 3.24 | 0.13 | 0 - 2 | 2 (11.11) | 0.23 | 0 - 2 | 0 (0) | 0.74 | 0 - 3 | 1 (5) | 0.131622 | 0.233719 |
| PQ4.9.57 Over past week, have you had unintentional weight changes? | Unintentional weight changes | 1.93 | 0.36 | 0 - 2 | 7 (38.89) | 0.91 | 0 - 3 | 0 (0) | 1.75 | 0 - 3 | 0 (0) | 0.182924 | 0.233719 |
| PQ4.9.18 Over past week, have you had back weakness? | Back weakness | 1.56 | 0.29 | 0 - 1 | 1 (5.56) | 1.09 | 0 - 3 | 0 (0) | 1.70 | 0 - 3 | 0 (0) | 0.233719 | 0.233719 |

**Table S3**. Pearson correlation coefficient matrix measuring correlations between pairs of variables from the Raman LDA model and potential confounders; correlations with values > 0.5 were highlighted.

|  |  | **A** | **B** | **C** | **D** | **E** | **F** | **G** | **H** | **I** | **J** | **K** | **L** | **M** | **N** | **O** | **P** | **Q** | **R** |
| --- | --- | --- | --- | --- | --- | --- | --- | --- | --- | --- | --- | --- | --- | --- | --- | --- | --- | --- | --- |
| **LD1** | **A** | **1** | -0.48 | -0.02 | 0.04 | 0.18 | -0.18 | 0.17 | -0.21 | -0.05 | -0.24 | -0.15 | 0.3 | -0.18 | -0.13 | -0.02 | -0.08 | -0.06 | 0.01 |
| **LD2** | **B** | -0.48 | **1** | -0.06 | -0.14 | -0.2 | 0.06 | -0.22 | 0.06 | -0.03 | -0.06 | 0.19 | -0.27 | 0.29 | 0.38 | 0.11 | 0.09 | 0.15 | 0 |
| **Medicine last 3 months (Y/N)** | **C** | -0.02 | -0.06 | **1** | **0.56** | 0.16 | 0.34 | 0.19 | 0.07 | 0.19 | 0.09 | 0.14 | 0.1 | -0.09 | -0.08 | -0.06 | 0.23 | 0.09 | 0.04 |
| **Medicine current (Y/N)** | **D** | 0.04 | -0.14 | **0.56** | **1** | 0.2 | 0.41 | 0.3 | 0.14 | 0.2 | 0.15 | 0.17 | 0.19 | -0.19 | -0.13 | -0.24 | 0.11 | 0.06 | -0.05 |
| **Supplement Count** | **E** | 0.18 | -0.2 | 0.16 | 0.2 | **1** | -0.14 | **0.64** | -0.15 | -0.26 | -0.2 | -0.11 | 0.09 | 0.02 | 0.07 | -0.34 | 0.09 | 0.02 | -0.21 |
| **Medication Count** | **F** | -0.18 | 0.06 | 0.34 | 0.41 | -0.14 | **1** | -0.19 | 0.41 | 0.32 | 0.49 | 0.39 | 0.36 | -0.13 | -0.02 | 0.14 | 0 | 0.04 | 0.07 |
| **Supplements Present** | **G** | 0.17 | -0.22 | 0.19 | 0.3 | **0.64** | -0.19 | **1** | -0.18 | -0.18 | -0.14 | -0.23 | -0.01 | 0.03 | -0.21 | -0.44 | -0.14 | 0.09 | -0.18 |
| **Medication_Class_Gabapentinoid_Present** | **H** | -0.21 | 0.06 | 0.07 | 0.14 | -0.15 | 0.41 | -0.18 | **1** | 0.27 | 0.42 | 0.24 | -0.08 | -0.16 | 0.1 | 0 | -0.05 | -0.11 | -0.07 |
| **Medication_Class_NSAID_Paracetamol_Present** | **I** | -0.05 | -0.03 | 0.19 | 0.2 | -0.26 | 0.32 | -0.18 | 0.27 | **1** | **0.59** | -0.01 | 0.02 | 0.05 | -0.11 | 0.2 | 0.16 | 0.02 | 0.12 |
| **Medication_Class_Opiate_Present** | **J** | -0.24 | -0.06 | 0.09 | 0.15 | -0.2 | 0.49 | -0.14 | 0.42 | **0.59** | **1** | 0.12 | -0.01 | 0 | -0.13 | 0.02 | 0.07 | -0.19 | -0.01 |
| **Medication_Class_SSRI_SNRI_Present** | **K** | -0.15 | 0.19 | 0.14 | 0.17 | -0.11 | 0.39 | -0.23 | 0.24 | -0.01 | 0.12 | **1** | -0.08 | -0.23 | 0.13 | 0.15 | -0.11 | -0.1 | 0.07 |
| **Medication Class Tricyclic Or Mirtazapine Present** | **L** | 0.3 | -0.27 | 0.1 | 0.19 | 0.09 | 0.36 | -0.01 | -0.08 | 0.02 | -0.01 | -0.08 | **1** | -0.18 | 0.11 | 0 | 0.13 | -0.01 | 0.17 |
| **Sex** | **M** | -0.18 | 0.29 | -0.09 | -0.19 | 0.02 | -0.13 | 0.03 | -0.16 | 0.05 | 0 | -0.23 | -0.18 | 1 | 0.02 | -0.07 | -0.01 | 0.25 | -0.13 |
| **Age at survey** | **N** | -0.13 | 0.38 | -0.08 | -0.13 | 0.07 | -0.02 | -0.21 | 0.1 | -0.11 | -0.13 | 0.13 | 0.11 | 0.02 | **1** | 0.04 | 0.37 | 0.06 | 0.14 |
| **BMI** | **O** | -0.02 | 0.11 | -0.06 | -0.24 | -0.34 | 0.14 | -0.44 | 0 | 0.2 | 0.02 | 0.15 | 0 | -0.07 | 0.04 | **1** | -0.03 | 0.04 | 0.34 |
| **Disease duration (years)** | **P** | -0.08 | 0.09 | 0.23 | 0.11 | 0.09 | 0 | -0.14 | -0.05 | 0.16 | 0.07 | -0.11 | 0.13 | -0.01 | 0.37 | -0.03 | **1** | 0.1 | -0.07 |
| **Processing duration (seconds)** | **Q** | -0.06 | 0.15 | 0.09 | 0.06 | 0.02 | 0.04 | 0.09 | -0.11 | 0.02 | -0.19 | -0.1 | -0.01 | 0.25 | 0.06 | 0.04 | 0.1 | **1** | 0.09 |
| **Recruiting time/freezing duration (days)** | **R** | 0.01 | 0 | 0.04 | -0.05 | -0.21 | 0.07 | -0.18 | -0.07 | 0.12 | -0.01 | 0.07 | 0.17 | -0.13 | 0.14 | 0.34 | -0.07 | 0.09 | **1** |

**Table S4.** Top Raman peak features selected based on descending LDA contribution. At each Raman peak, averaged quantification of HC, ME and MS is shown. Statistical comparison is shown as p value by a global student t-test.

| **Raman Peak (cm^–1^)** | **LDA contribution** | **HC average** | **ME average** | **MS average** | **P value** |
| --- | --- | --- | --- | --- | --- |
| **467.324** | 285.43 | 0.212 | 0.201 | 0.109 | 3.42E-10 |
| **1180.3** | 279.03 | 1.501 | 1.458 | 1.506 | 0.03603478 |
| **1366.86** | 276.80 | 2.203 | 2.242 | 2.34 | 1.02E-05 |
| **1173.68** | 255.38 | 1.622 | 1.59 | 1.663 | 0.0226378 |
| **742.745** | 255.23 | 1.374 | 1.35 | 1.462 | 0.0016585 |
| **3054.15** | 253.30 | 1.258 | 1.261 | 1.202 | 0.00018715 |
| **2810.91** | 237.08 | 0.256 | 0.28 | 0.278 | 0.11974009 |
| **520.949** | 231.16 | 0.487 | 0.495 | 0.41 | 2.85E-05 |
| **1631.46** | 220.60 | 1.13 | 1.128 | 1.095 | 0.45513773 |
| **431.398** | 218.41 | 0.448 | 0.432 | 0.3 | 8.01E-10 |
| **1347.41** | 191.05 | 2.37 | 2.434 | 2.505 | 9.06E-07 |
| **460.15** | 186.97 | 0.254 | 0.223 | 0.137 | 9.73E-10 |
| **1080.47** | 186.61 | 1.922 | 1.907 | 1.953 | 0.12243916 |
| **1700.24** | 179.91 | 0.656 | 0.632 | 0.525 | 0.00016772 |
| **1482.64** | 179.09 | 1.525 | 1.565 | 1.642 | 0.00402072 |
| **1587.41** | 175.63 | 1.196 | 1.235 | 1.247 | 0.32159906 |
| **3030.43** | 171.42 | 1.167 | 1.149 | 1.109 | 0.00370214 |
| **556.523** | 165.11 | 0.509 | 0.539 | 0.448 | 0.00016625 |
| **3062.04** | 155.37 | 1.189 | 1.195 | 1.135 | 0.00012515 |
| **348.223** | 155.31 | 0.823 | 0.785 | 0.701 | 1.55E-05 |
| **1130.53** | 154.44 | 1.583 | 1.56 | 1.642 | 0.13577574 |
| **395.327** | 152.81 | 0.603 | 0.587 | 0.443 | 1.19E-06 |
| **416.986** | 150.81 | 0.639 | 0.622 | 0.481 | 3.13E-08 |
| **2824.48** | 146.38 | 0.474 | 0.496 | 0.518 | 0.05868149 |
| **373.617** | 142.63 | 0.579 | 0.548 | 0.427 | 1.30E-07 |
| **1023.39** | 142.23 | 1.588 | 1.543 | 1.564 | 0.0402755 |
| **794.753** | 140.72 | 1.142 | 1.128 | 1.09 | 0.05205117 |
| **1389.5** | 137.58 | 1.519 | 1.551 | 1.596 | 0.03224369 |
| **2859.63** | 132.97 | 3.523 | 3.545 | 3.742 | 0.00236603 |
| **1536.77** | 128.30 | 0.572 | 0.582 | 0.588 | 0.91035318 |
| **770.519** | 119.92 | 1.382 | 1.357 | 1.323 | 0.11458769 |
| **767.054** | 118.03 | 1.157 | 1.138 | 1.093 | 0.00822416 |
| **1489.03** | 116.36 | 1.155 | 1.172 | 1.22 | 0.20345991 |
| **577.799** | 115.99 | 0.374 | 0.375 | 0.293 | 0.0001191 |
| **2851.53** | 114.21 | 2.692 | 2.7 | 2.88 | 0.00628556 |
| **1408.85** | 112.36 | 1.454 | 1.477 | 1.492 | 0.20159483 |
| **2988.08** | 109.69 | 4.008 | 4.047 | 4.061 | 0.5193555 |
| **2800.04** | 109.02 | 0.19 | 0.202 | 0.2 | 0.66904405 |
| **427.797** | 108.79 | 0.497 | 0.478 | 0.342 | 3.48E-10 |
| **972.718** | 108.42 | 1.602 | 1.641 | 1.575 | 0.12771077 |
| **3056.78** | 107.30 | 1.262 | 1.281 | 1.222 | 0.00011284 |
| **495.963** | 107.08 | 0.474 | 0.471 | 0.411 | 0.00162004 |
| **1428.16** | 106.76 | 1.874 | 1.901 | 1.978 | 7.74E-05 |
| **1026.76** | 103.74 | 1.741 | 1.705 | 1.704 | 0.1179658 |
| **1524.07** | 103.61 | 0.777 | 0.739 | 0.806 | 0.34013404 |
| **753.17** | 103.19 | 1.102 | 1.103 | 1.095 | 0.90952865 |
| **1305.12** | 100.76 | 3.103 | 3.121 | 3.28 | 2.20E-10 |
| **894.445** | 98.81 | 1.446 | 1.415 | 1.333 | 2.35E-06 |
| **1593.72** | 97.72 | 1.04 | 1.058 | 1.031 | 0.62109426 |
| **1590.56** | 96.37 | 1.081 | 1.111 | 1.089 | 0.50027145 |
| **818.92** | 96.03 | 1.213 | 1.18 | 1.116 | 6.12E-07 |
| **641.33** | 95.49 | 0.595 | 0.614 | 0.554 | 0.00220252 |
| **1040.22** | 95.30 | 1.677 | 1.626 | 1.521 | 3.12E-05 |
| **1450.64** | 92.45 | 2.876 | 2.911 | 3.041 | 3.00E-11 |
| **1331.17** | 91.13 | 3.259 | 3.295 | 3.443 | 1.35E-06 |
| **2854.23** | 88.61 | 2.912 | 2.931 | 3.124 | 0.0015022 |
| **860.195** | 88.55 | 1.409 | 1.357 | 1.287 | 7.35E-09 |
| **965.94** | 87.58 | 1.642 | 1.666 | 1.598 | 0.16692987 |
| **1672.17** | 86.37 | 1.748 | 1.795 | 1.823 | 0.02620277 |
| **798.208** | 84.25 | 1.09 | 1.054 | 1.002 | 1.37E-05 |
| **1160.43** | 84.21 | 1.538 | 1.471 | 1.542 | 0.05395872 |
| **2958.83** | 84.11 | 7.167 | 7.298 | 7.484 | 0.01510187 |
| **3022.51** | 83.74 | 1.323 | 1.32 | 1.288 | 0.08256413 |
| **1759.18** | 82.15 | 0.23 | 0.078 | -0.131 | 5.44E-12 |
| **1469.85** | 81.14 | 1.886 | 1.914 | 1.977 | 0.0026206 |
| **1301.86** | 80.60 | 3.081 | 3.086 | 3.241 | 1.22E-10 |
| **711.395** | 80.05 | 0.702 | 0.691 | 0.634 | 0.00123915 |
| **1090.51** | 79.61 | 2.097 | 2.09 | 2.15 | 0.03577563 |
| **1584.25** | 79.09 | 1.396 | 1.424 | 1.47 | 0.23850419 |
| **1555.8** | 75.19 | 0.866 | 0.886 | 0.88 | 0.70012075 |
| **669.421** | 75.05 | 0.675 | 0.664 | 0.617 | 0.00490924 |
| **2993.39** | 75.01 | 3.313 | 3.347 | 3.323 | 0.49207105 |
| **488.812** | 74.82 | 0.522 | 0.505 | 0.447 | 0.00089672 |
| **506.679** | 74.12 | 0.356 | 0.35 | 0.253 | 1.74E-08 |
| **2865.03** | 73.20 | 4.366 | 4.408 | 4.662 | 0.00030988 |
| **1476.25** | 71.44 | 1.729 | 1.761 | 1.832 | 0.00564951 |

**Table S5.** Model performance in accuracy by individual classifiers from the ensemble learner. LDA: Linear Discriminant Analysis; XGB: Extreme Gradient Boosting; SVM-Linear: Support Vector Machine with Linear Kernel: SVM-Radial: Support Vector Machine with Radial Basis Function Kernel; MLPNN: Monotone Multi-Layer Perceptron Neural Network; RF: Random Forest; MDA: Mixture Discriminant Analysis; GBM: Stochastic Gradient Boosting.

| ***Classifier*** | ***Hyperparameters*** | ***Performance on train set*** | ***Performance on test set*** |
| --- | --- | --- | --- |
| **LDA** | **dimension = 4** | **54.8%** | **47.1%** |
| **XGB** | **nrounds = 150, max_depth = 3, eta = 0.4, gamma = 0, subsample = 1, colsample_bytree = 0.8, rate_drop**  **= 0.01, skip_drop = 0.95 and min_child_weight = 1** | **84.3%** | **51.2%** |
| **SVM-Linear** | **Cost = 1** | **68.4%** | **48.8%** |
| **SVM-Radial** | **mtry = 100, splitrule = extratrees, min.node.size = 1** | **60.1%** | **49.9%** |
| **MLPNN** | **hidden1 = 15, n.ensemble = 5** | **82.6%** | **50.2%** |
| **RF** | **mtry = 109, splitrule = extratrees, min.node.size = 1** | **87.4%** | **61.2%** |
| **MDA** | **subclasses = 4** | **60.1%** | **53.1%** |
| **GBM** | **n.trees = 150, interaction.depth = 3, shrinkage = 0.1, n.minobsinnode = 10.** | **80.9%** | **48.1%** |
